# Divergent functional connectivity changes associated with white matter hyperintensities

**DOI:** 10.1101/2023.03.05.23286807

**Authors:** Alexander F. Santillo, Tor O. Strandberg, Nina H. Reislev, Markus Nilsson, Erik Stomrud, Nicola Spotorno, Danielle van Westen, Oskar Hansson

## Abstract

Age-related white matter hyperintensities are a common feature and known to negatively impact structural integrity, functional connectivity and cognitive performance. However, this has yet to be fully understood mechanistically. We analysed multiple MRI modalities acquired in 465 non-demented individuals from the Swedish BioFINDER study including 334 cognitively normal and 131 participants with mild cognitive impairment. White matter hyperintensities were automatically quantified using fluid-attenuated inversion recovery MRI and parameters from diffusion tensor imaging were estimated in major white matter fibre tracts. We calculated fMRI resting state-derived functional connectivity within and between predefined cortical regions structurally linked by the white matter tracts and evaluated how change in functional connectivity is affected by white matter lesions and related to cognition, in the form of executive function and processing speed. We examined the functional changes using a measure of sample entropy. As expected hyperintensities were associated with disrupted structural white matter integrity and were linked to reduced functional *interregional* lobar connectivity, which related to decreased processing speed and executive function. Simultaneously, hyperintensities were also associated with increased *intraregional* functional connectivity, but only within the frontal lobe. This phenomenon was also associated with reduced cognitive performance. The increased connectivity was linked to increased entropy (reduced predictability and increased complexity) of the involved voxels’ blood oxygenation level dependent signal. Our findings expand our previous understanding of the impact of white matter hyperintensities on cognition by describing novel mechanisms that may be important beyond this particular type of brain lesions.

## 1. Introduction

Brain white matter hyperintensities (WMHs) are associated with long-standing hypertension and arteriosclerosis (van der Flier et al. 2018; Debette and Markus 2010). Increased WMH volume is related to cognitive decline (Prins and Scheltens 2015; Kandel et al. 2016; Wang et al. 2017). Thanks to the developments in neuroimaging our understanding of how this relationship is mediated has increased considerably. Importantly a link between WHM, white matter integrity as assessed using diffusion MRI, and brain function as assessed by functional MRI has been established (Langen et al. 2017, Reijmer et al. 2015, Tuladhar et al. 2015). However, only some facets of brain functionality have hitherto been assessed, and the mechanism with which WMHs disrupt functional networks and lead to the reduction in cognitive performance remain unexplained.

Prefrontal white matter is often preferentially affected by WMHs and lesions in this region are commonly associated with deficits in executive, “prefrontal” function (Duering et al. 2014). However, a number of studies provided impetus to revise this locationalist view. Tullberg et al. (Tullberg et al 2004) showed that any cerebral location of WMH was associated with frontal hypometabolism and executive deficits. Similarly, Jacobs et al. (Jacobs et al. 2012) showed that hyperintensities not only in frontal, but also parietal white matter, are important mediators of decline in executive function.

Assessing white matter involvement outside the visually apparent WHMs was therefore seen as potentially informative (O’Sullivan et al. 2001; Catani and Ffytche 2005; Lawrence et al. 2018). The structural white matter network can be quantified using diffusion MRI, which provides information on microstructural aspects of white matter (Beaulieu 2002). The technique has revealed decreased fractional anisotropy (FA) and increased mean diffusivity (MD) within WMHs, but also in normal appearing white matter, seemingly related to cognition (Tuladhar et al. 2015). Tuladhar and coworkers (Tuladhar et al. 2015) showed that severity of small vessel disease was associated not only with diffusion tensor metrics, but also with parameters from tractography-derived graph theoretical analysis.

Adding functional to structural connectivity became an obvious step further. In a cognitively normal group with WMHs, Reijmer and coworkers (Reijmer et al. 2015) found that reduced default mode resting state network connectivity was related to altered diffusion parameters of the cingulum bundle - a central structural link of that functional network. Very similar findings have been reported for another structural link in the default mode network - the inferior fronto-occipital fasciculus (IFOF) (Taylor et al. 2016). Langen and coworkers (Langen et al. 2017) examined the relation between functional connectivity and WMHs in tracts directly linking functional subunits, but also how lesions impacted indirect (no direct anatomical link) connections. They found that tract-specific WMHs resulted in reduced functional connectivity between both cortical regions with direct anatomical links as well as those indirectly linked. This likely reflects the network-based organisation of the brains’ communication and its functional response caused by insults to its structural network integrity.

In this study we sought to expand on existing knowledge. Particularly we wanted to a) examine to what extent the impact of WMHs on the link between structural and functional connectivity affect cognition and b) to examine how functional connectivity can be interpreted using signal complexity measures. To this end, we mapped out functional connectivity changes associated with WMH-related insults to the structural integrity of central tracts. We applied a combination of WMH volume, diffusion MRI, and resting-state fMRI, further relating our findings an entropy-based measure of information content (Richman and Moorman 2000). Since concomitant amyloid pathology and WMH is common and it is a matter of debate if and how they interact towards cognitive impairment (Marchant et al 2011, Taylor et al 2016), we also studied the potential effects of amyloid pathology.

## 2. Material and Methods

### 2.1 Cohort

Our study population is part of the prospective Swedish BioFINDER Study (NTC012086675) and recruited between 2010 and 2015. The 465 participants were classified as 1) cognitively normal (CN; n=334) (Jack et al. 2018) of which 118 had subjective cognitive decline (Mattsson et al. 2016), or 2) having mild cognitive impairment (MCI; n=131) (Petersen 2007). Study design and specific inclusion/exclusion criteria are described in the Supplementary Material (see also http://biofinder.se).

Demographics are shown in Table 1. Amyloid pathology was assessed using the CSF Aβ42/Aβ40 ratio (Janelidze et al. 2016) with a cut-off of 0.1.

**Table 1:**
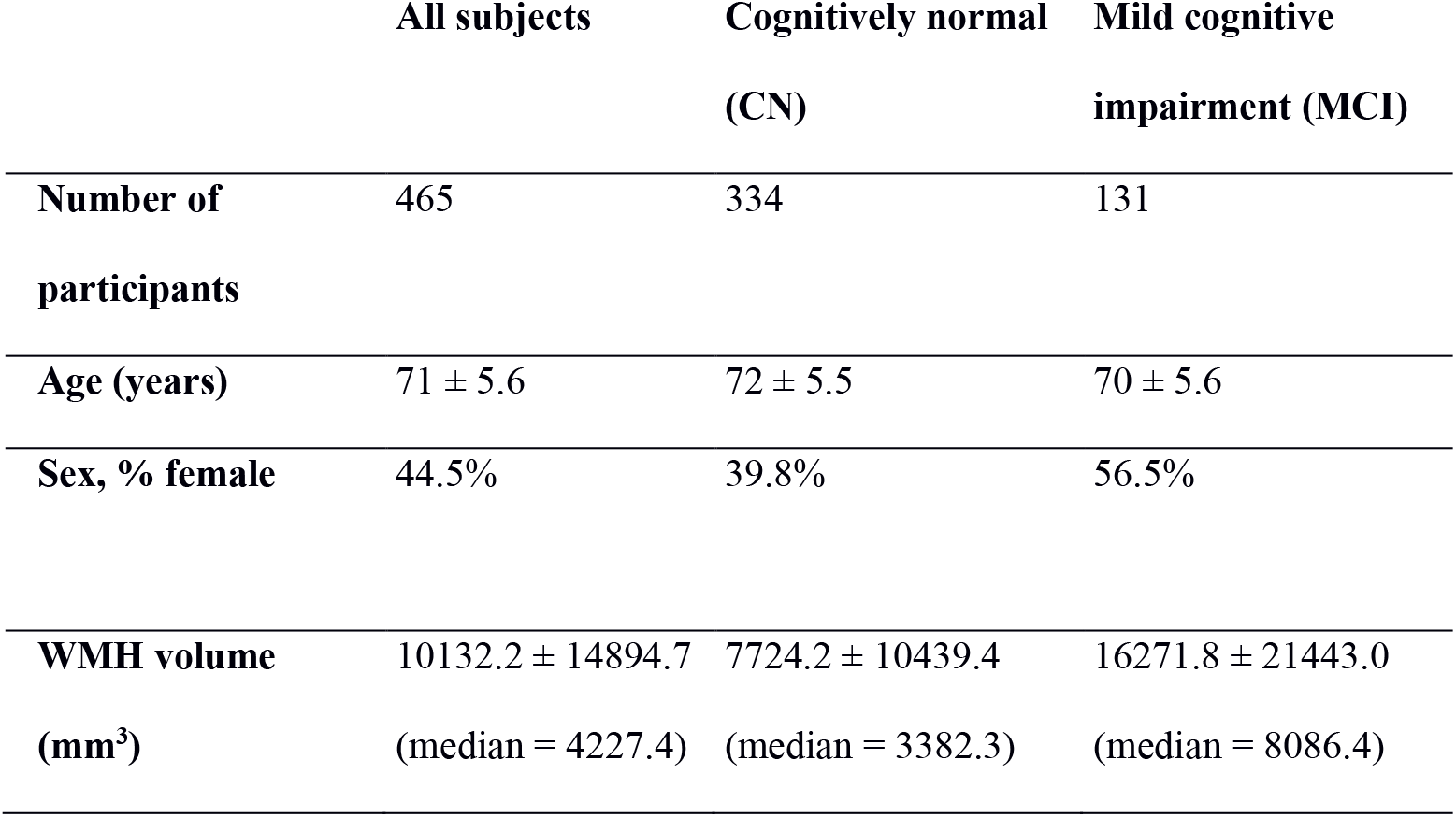
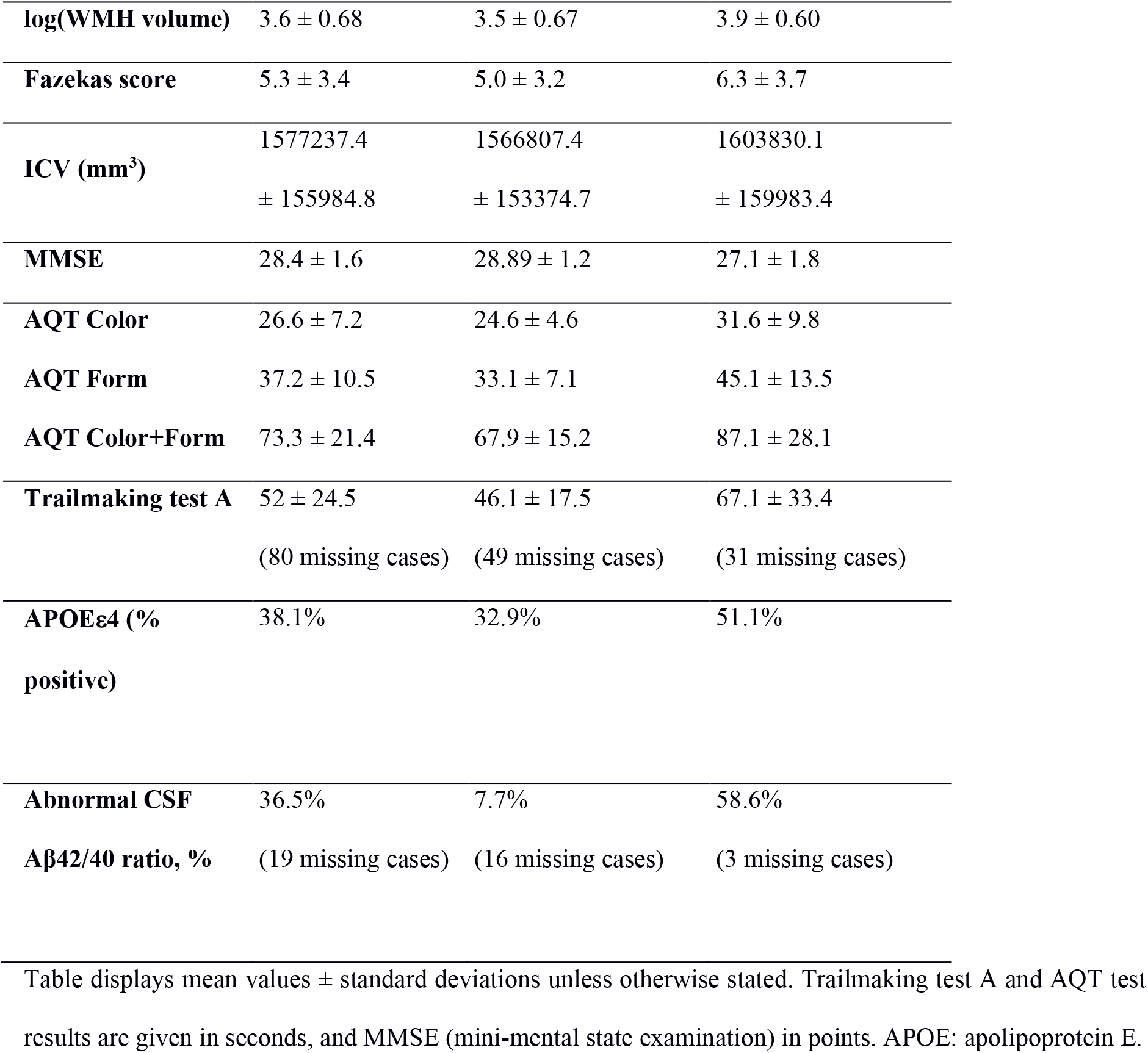
Cohort characteristics.

### 2.2 Image acquisition and preprocessing of imaging data

Details on image acquisition parameters and preprocessing of resting-state fMRI data are provided in the Supplementary Material. In short, imaging was performed on a 3.0T Siemens Tim Trio scanner (Siemens Medical Solutions, Erlangen, Germany). Structural 3D T1-weighted MPRAGE (1×1×1.2 mm^3^) and T2-weighted FLAIR (0.7×0.7×5.2 mm^3^) images were acquired for anatomical information and detection of WMHs. Resting-state fMRI (eyes closed, 3×3×3 mm^3^, 180 dynamic scans, 6 minutes) was performed for assessment of functional connectivity and diffusion-weighted MRI (64 diffusion encoding directions, b-values of 0 and 1000 s/mm^2^, 2×2×2 mm^3^) for diffusion tensor imaging.

Resting-state fMRI data were preprocessed using FMRIB Software Library (FSL) (Jenkinson et al. 2012), Analysis of Functional NeuroImage (AFNI) (Cox 1996), and Advanced Normalization Tools (ANTs) (Avants et al. 2014). Anatomical processing of the T1-weighted image involved skull stripping, segmentation of white matter, grey matter and CSF and normalization to 1×1×1 mm^3^ MNI152-template space (Grabner et al. 2006) with the non-linear diffeomorphic mapping implemented in ANTs. Dropping the first five frames in anticipation of steady state, functional data was bulk motion and slice timing corrected, furthermore nuisance regressed using the white matter/CSF average signal, 6 components of physiological noise (Behzadi et al. 2007), 24 motion parameters (Friston et al. 1996), and linear/quadratic trends. Finally, the functional data were transformed to MNI-space using the T1 non-linear normalization warp. The diffusion-weighted images were corrected for motion and eddy current induced artefacts by registering all images to the first non-diffusion weighted image, using Elastix (Nilsson et al. 2015). Maps of MD and FA were calculated from the diffusion tensor eigenvalues using in-house developed software implemented in Matlab (MATLAB 2013a, The MathWorks Inc., Natick, MA, USA). All MD and FA maps were visually inspected for artefacts.

### 2.3 Definition of fibre tracts of interest

In our analysis we employed the John Hopkins University (JHU) tractography atlas (in MNI template space), containing probabilistic templates of fibre tracts (Mori et al. 2005). The atlas provides the following 10 fibre tracts in the left and right hemisphere: forceps major, forceps minor, uncinate fasciculus (UF), inferior longitudinal fasciculus (ILF), inferior fronto-occipital fasciculus (IFOF), superior longitudinal fasciculus (SLF), cingulum hippocampal part (Cing-hipp), cingulum cingulate gyrus part (Cing-cing), cortico-spinal tract (CST), and anterior thalamic radiation (ATR). CST and SLF were discarded in the analyses due to difficulty in defining suitable endpoint regions for the subsequent resting-state analysis. The tracts were used in their full probabilistic form and additionally as binary mask based on a 10% threshold of the tract probability.

### 2.4 Assessment of white matter hyperintensities

The Lesion Segmentation Tool (LST) v. 1.2.3 implemented in SPM 5 (Schmidt et al. 2012) was used for automated segmentation of lesions, based on the T1-weighted and FLAIR images with the parameter kappa set to 0.4 (further details on lesion definition given in Supplementary Methods). To evaluate WMH volume (mm^3^) a 25% threshold on the probabilistic LST maps and a cluster filter requiring a minimum of 10 constituent voxels were applied. The WMH maps were transformed to MNI space using the non-linear transform extracted in normalizing the high-resolution T1-weighted image. Also, the amount of WMHs were scored using the Fazekas scale (Fazekas et al. 1993).

### 2.5 Assessment of MD and FA within fibre tracts

The MD and FA maps were rigidly coregistered to each subject’s T1-weighted images with ANTs and then warped to MNI space. Subject- and tract-specific weighted averages of MD and FA were calculated according to the formula: *w*(*x*) = *m*(*x*)*p*_*tract*_(*x*)*p*_*WMH*_(*x*), where the weighted value *w* is formed by the measure *m*(*x*) (MD or FA) times the WMH and tract probabilities at voxel location *x*. Weighted MD and FA were implemented to more accurately account for combined lesion and tract presence than corresponding binarized probability masks. To assess the impact of WMHs on the fibre tracts for each subject, we calculated weighted average over all voxels for MD and FA: ∑ *w*(*x*)/∑ *p*_*tract*_(*x*)*p*_*WMH*_(*x*).

### 2.6 Functional connectivity analyses

All functional connectivity analyses were performed in terms of predefined A and B cortical region pairs connected by specific tracts outlined above (Supplementary Table 1). Rather than smoothing rs-fMRI data to reduce individual variability effects, we downsampled the processed blood oxygenation level dependent (BOLD) time series in template space to 6×6×6 mm^3^ resolution using trilinear interpolation. For each tract, voxel-wise connectivity maps were constructed by calculating Pearson correlations between all voxel pairs in the joint set spanned by A and B (correlations were variance stabilized via a Fisher-Z transform prior to statistical analysis). Voxel-wise connectivity maps spanning all voxels in pooled A and B regions were explored in full by calculating network components correlating with the corresponding tract damage metric (see Statistical Analysis subsection for details on algorithm). The tract damage metric was chosen as the WMH and tract probability-weighted MD sum defined above. The rationale for including MD information was to better account for WMH *severity*, as a large WMH volume is not always equivalent to a high severity of tissue damage, although this is often the case.

In order to clarify the nature of large-scale neuronal signal changes implicated in the functional connectivity analysis, we additionally analysed the information content in the related BOLD time series using an entropy measure for discrete time series – sample entropy (Richman and Moorman 2000). Embedding dimension was set to m=2 and the range commensurate with data time series standard deviation. The sample entropy is an information-theoretical measure which increases for time series with lower predictability and more random behaviour, and conversely decreases for a more ordered and predictable time series with less complexity. For each subject, we calculated the entropy for each voxel part of the calculated network component. The components total connectivity sum was then Spearman correlated with voxel entropy values to reveal the nature of the signal changes underlying the significant component (Bonferroni-corrected for the number of voxels at p<0.001).

### 2.7 Cognitive testing

We employed tests representing cognitive domains generally influenced by white matter lesions. A Quick Test for Cognitive Speed (AQT) consists of three subparts (AQT Color, AQT Form, and AQT Color and Form), were the first two subparts evaluate attention and psychomotor speed while AQT Color and Form additionally demand executive functions such as working memory and inhibition (Wiig et al. 2002). Additionally, Trailmaking test A provided information on visual scanning and motor speed (Reitan and Wolfson 1985).

### 2.8 Statistical analysis

WMH volumes were generally non-normally distributed and therefore log-transformed prior to analysis. The associations between total WMH volume and age, gender, Fazekas score, and total intra-cranial volume, were tested using Pearson correlations (point-biserial for gender), while Spearman correlations were used between lesion loads in the different tracts (and global WMH volume). Bonferroni-corrected paired and unpaired t-tests was used to test for significant differences between intra- and extralesional measures. Functional connectivity changes associated with the tract damage metric (tract and lesion probability-weighted MD sum) were calculated as network components (similar to approach in (Zalesky et al. 2010) over all voxels of the joint set of the two structurally linked GM regions. Prior to component calculations, age and intercranial volume (ICV) were corrected for by means of partial correlation. The employed algorithm proceeds by calculating the largest component (defined as a connected set of links) for which *r*_*ij*_ *> r*_0_ (or *r*_*ij*_ < *r*_0_ for negative correlations), where *r*_*ij*_ is the Spearman correlation between connectivity *c*_*ij*_ (linking voxel *i* and *j*) and the tract damage metric. The initial correlation threshold *r*_0_ on every link was chosen such that *p*_*ij*_ < 0.001 (uncorrected) for all links candidating for the network component. The component size was defined as the number of constituent links multiplied by the Spearman correlation of tract damage metric and the components’ connectivity sum. To correct for multiple comparisons, we compared the size of the resulting component with a permutation-generated null distribution of component sizes. By repeatedly randomizing connectivity and damage metric data pairs and seeking the largest resulting component, the construction of a null distribution of sizes allowed assignment of a corrected component p-value. The randomization process was repeated a large number of times until the resulting p-value converged, whereby significance was declared if *p* < 0.05, thus controlling for the family-wise error at *α* = 0.05. We then studied the characteristics of the significant network components by summing the connectivity of constituent links and relating them to other variables of interest, using Spearman correlations.

### 2.9 Ethics and data availability

The Regional Ethics Committee in Lund, Sweden, gave ethical approval of the study design and all subjects gave their written informed consent in accordance with the Declaration of Helsinki. Anonymized data will be shared by request from a qualified academic investigator for the sole purpose of replicating procedures and results presented in the article if data transfer is in agreement with EU legislation on the general data protection regulation and decisions by the Ethical Review Board of Sweden and Region Skåne, which should be regulated in a material transfer agreement.

## 3. Results

### 3.1 Tract vulnerability to WMHs

Whole-brain WMH volume correlated significantly with age (*r=*0.35, p<0.001) and ICV (*r=*0.30, p<0.001), hence both were considered as covariates in the network component calculations. The frequency map of whole-brain WMHs revealed a clear periventricular pattern extending deep into the white matter (see Fig. 1A). Fig. 1B illustrates the correlation between tract damage metric for different tracts, providing information on lesion distribution within tracts and lesion correlation among tracts. Evaluating the distribution of WMHs across tracts, (combining 10% threshold on tract probability and 25% lesion probability) IFOF, forceps major and ATR exhibited the highest total WMH volumes (see Fig. 1C). The ILF, UF and forceps minor also had a considerable amount of hyperintensities, while the cingulum was relatively unaffected. Tract-wise sums of lesion volumes might not account for the relatively higher intersubject spatial variability of some tracts. Fig 1D illustrates the tract’s focality of lesion damage.

**Fig. 1:**
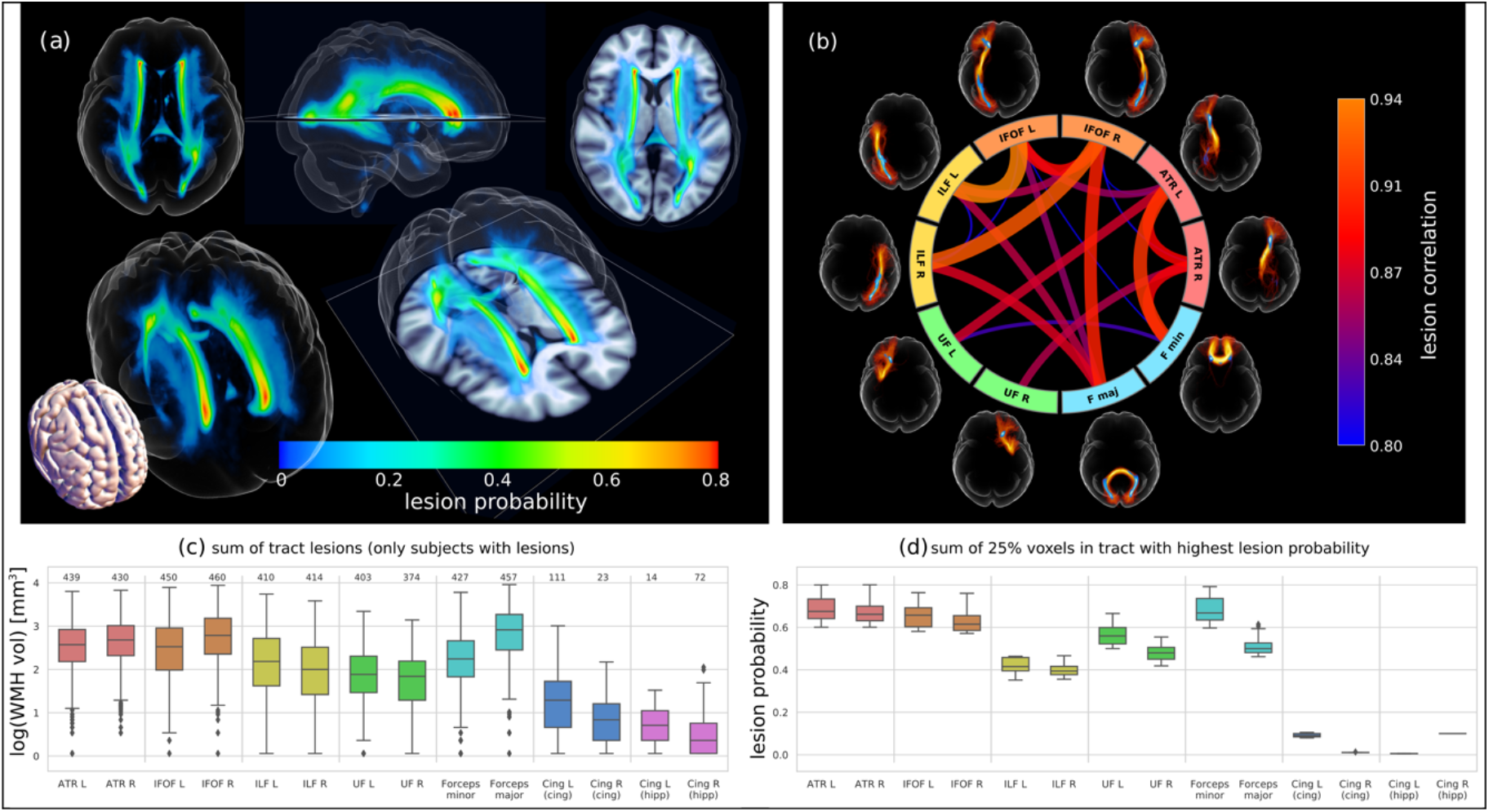
Lesion probabilities. Panel (a) shows maximal intensity projections of the lesion frequency map formed by all subjects with lesions. (b) shows the correlation of lesion loads between the different tracts, revealing for example that the IFOF L correlates highest with concomitant ILF L load. In (c), boxplots show the distribution of log-transformed total WMH volume on the tracts of interest. (d) shows a measure of focality; the lesion frequency map is masked with >10% tract probability and the top 25% of voxel frequency values are summed, illustrating that the topology of the damage profiles. Note that Forceps major has a rather high lesion volume (see c) but a much more diffuse damage profile.

### 3.2 Microstructural properties of the tracts

Fig. 2A shows the probability-weighted MD average for each tract, comparing whole tract (for subjects with no lesions in tract), extra- and intralesional values (for subjects with non-zero metric). The analogous plot for FA is shown in Fig. 2B. Whereas the MD averages serve to evaluate qualitative tissue changes, the effective communication disruption should be more accurately gauged by the total tract load, i.e. the intralesional probability-weighted MD sum used in the connectivity calculations (see Fig. 2C).

**Figure 2:**
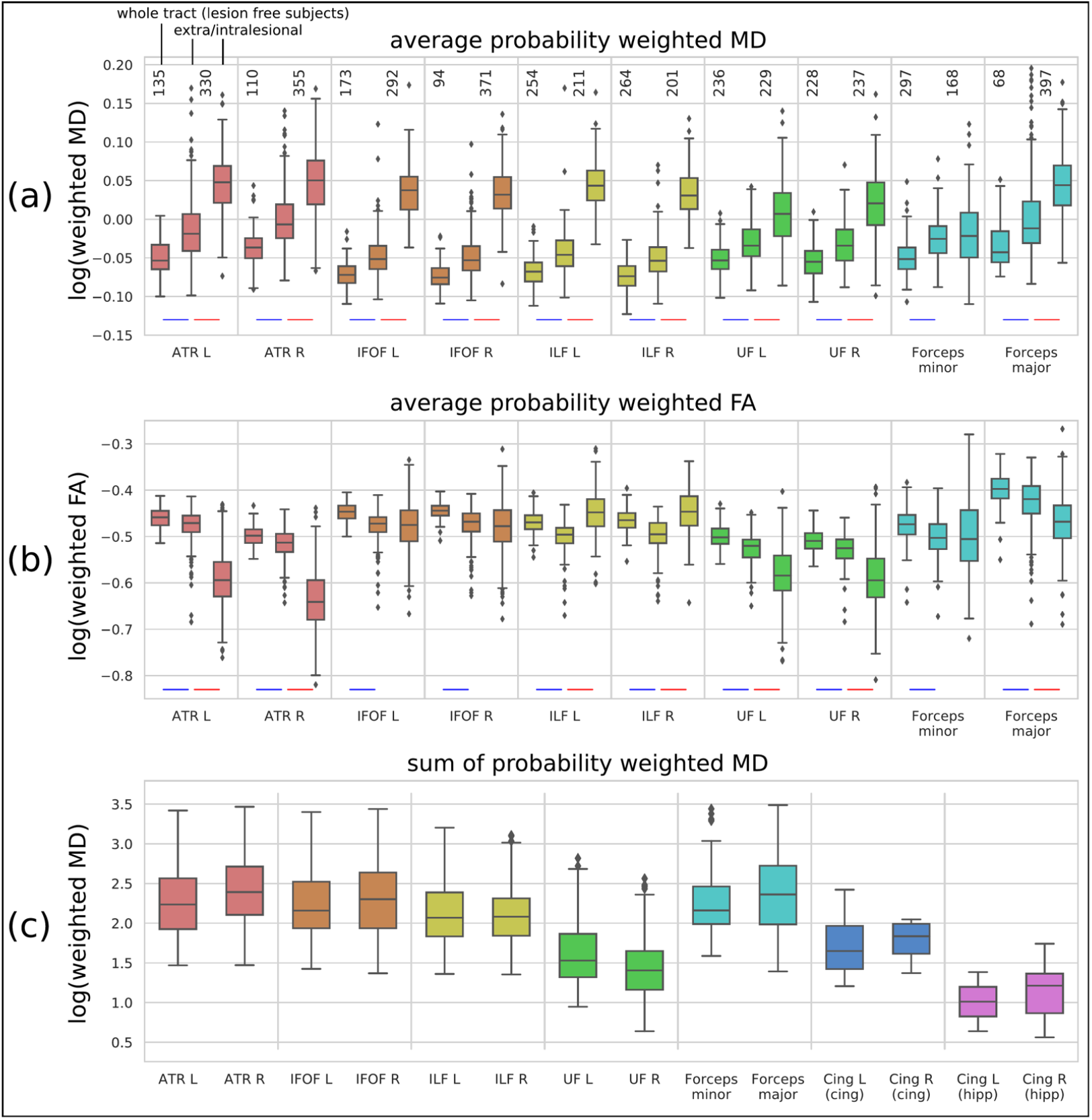
Altered fiber bundle topology mirrored in MD and FA statistics. (a) shows how the probability weighted MD average varies across the whole tract values for subjects with no lesions on tract, followed by the extra- and intralesional values. Number of subjects in boxplot statistics is shown above boxes. Blue and red line under boxes indicate Bonferroni corrected significance (p<0.001) in unpaired and paired t-test, respectively. (b) show analogous weighted statistics for FA and (c) the intralesional MD sum per tract, which quantifies the total tract damage and is used in the connectivity calculations.

The DTI metrics of the tracts were clearly affected by WMHs, evident in the relative MD increase and FA decrease of intra-versus extralesional averages. Most tracts revealed a statistically significant difference (p<0.001) between intra- and extralesional MD, except forceps minor and cingulum. Whole tract averages in subjects with no lesions were significantly different from extralesional values of subjects with lesions (p<0.001), in all tracts. This shows that even beyond the detectable WMH hotspots, extralesional changes are consistently present, albeit with smaller magnitudes. Corresponding statistics in FA were less consistent with fewer tracts showing significant differences. Due to the recorded difference in reliability between the two measures (see discussion), we focused on MD as the more adequate probe for altered topology.

### 3.3 The effect of regional WMHs on functional connectivity

For each tract we calculated functional connectivity patterns correlating with the tract damage metric (tract and lesion probability-weighted MD sum) across all subjects. The resulting network component was computed from all links within and between the two grey matter regions connected by the tract. Supplementary Table 2 provides an overview of the correlation values between the tract damage metric and the summed connectivity on significantly correlating network components (positive and negative correlations).

The tracts with the largest lesion burden, i.e. IFOF and ATR, yielded significant network components of both increased and decreased connectivity with increasing damage metric. (see Fig. 3 and 4 and Supplementary Videos 1 to 4). Clear qualitative patterns were associated with the tracts’ damage metric, consisting of reduced connectivity between the two structurally linked grey matter regions (Fig. 3B/E and 4B/E) and a concomitantly increased intraregional connectivity anteriorly (Fig3C/F and 4C/F). Whereas no great differences in tract damage profile (Fig. 3A and D) between IFOF left and right were apparent, ATR right had a distinctly higher tract times lesion probability maximum than the left counterpart (56% versus 28%), also seen in the reduced connectivity density (Fig. 4A/B and Fig. 4D/E). Similarly, decreased tract damage metric of ILF and left UF were also associated with decreased interregional connectivity (Supplementary Fig. 1 and 2). However, only tracts connecting to the frontal lobe had associations with increased functional connectivity (similar to IFOF and ATR) in the frontal region. Forceps minor damage was associated with a large interhemispherical network component of increased connectivity with laterality corresponding to structural damages (see Supplementary Fig. 3, no significant decrease found). Fig. 5 shows the regional voxelwise entropy correlations with the connectivity sum on the calculated component for forceps minor (Fig. 5AB) and ATR R (Fig. 5CD). These two components exhibited the highest correlations with entropy (see Supplementary Table 3 for all tract summary), indicating BOLD signal oxygenation patterns of growing complexity with increased connectivity. Note the entropy’s lateral correspondence for forceps minor (c.f. Figs. 5A and Supplementary Fig. 3). These results encircle the frontal increase in connectivity as related to less predictable oxygenation time series.

**Fig. 3:**
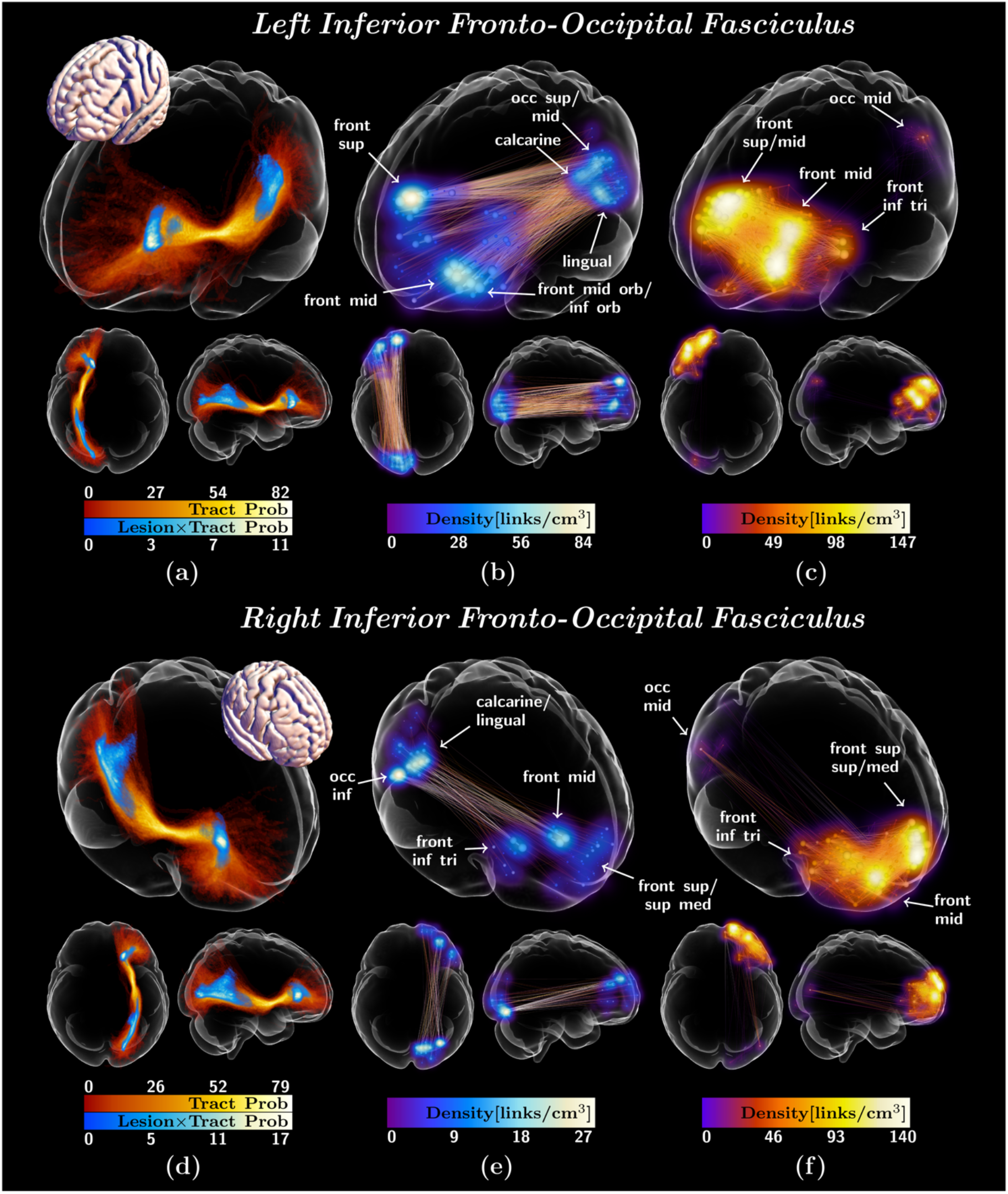
Tract damage and decreased/increased functional connectivity for left and right inferior fronto-occipital fasciculus (IFOF). Panel (a) and (d) show maximal intensity projections of the tract probability (warm colors) with overall lesion probability multiplied by tract probability overlaid (cold colors). (b)/(e) and (c)/(f) show the decrease/increase in functional connectivity (cold/warm colors used for density of links converging on voxels in grey matter region) associated with the tract damage metric for left and right IFOF, respectively. See also Supplementary Video 1 and 2. In the functional connectivity link plots of the significantly correlating network components, link density [links/cm^3^] has been approximated by convoluting with a Gaussian kernel and rendering as maximal intensity projections. Functional connectivity link endpoint transparencies and sphere sizes are proportional to the number of links converging on the corresponding voxel (to ensure link visibility maximal sphere size/link opacity set to 75th percentile plus 1.5 times interquartile range over all voxels’ link count).

**Fig. 4:**
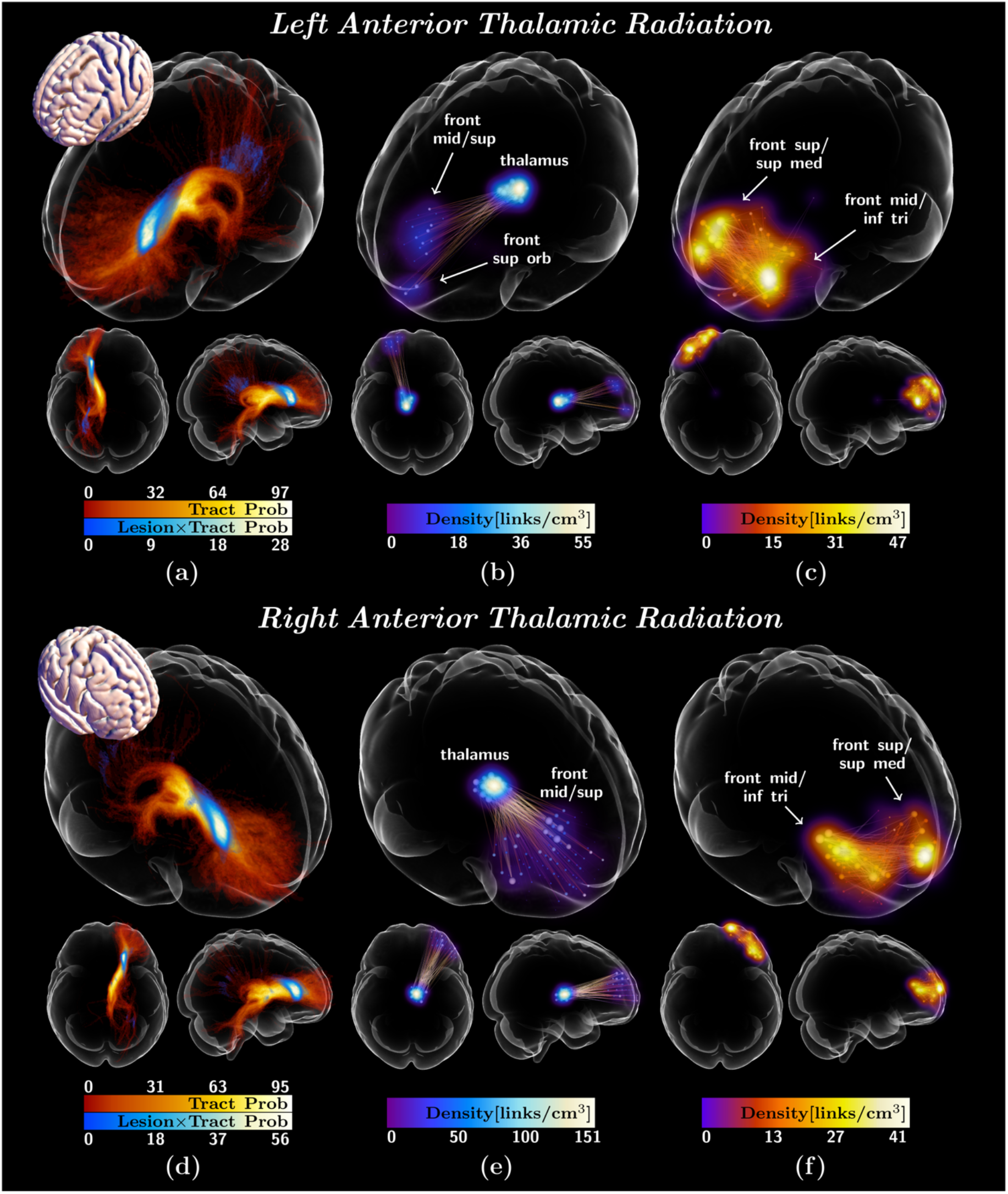
Tract damage overview and decreased/increased functional connectivity for left and right anterior thalamic radiation (ATR). (a) and (d) show maximal intensity projections of the tract probability (warm colors) and superimposed overall lesion probability multiplied by tract probability (cold colors). (b)/(c) and (e)/(f) show decreased/increased functional connectivity associated the with tract damage metric (cold/warm colors) for left and right ATR, respectively. See also Supplementary Video 3 and 4. See Figure 3 caption for details of the visualizations.

**Fig. 5:**
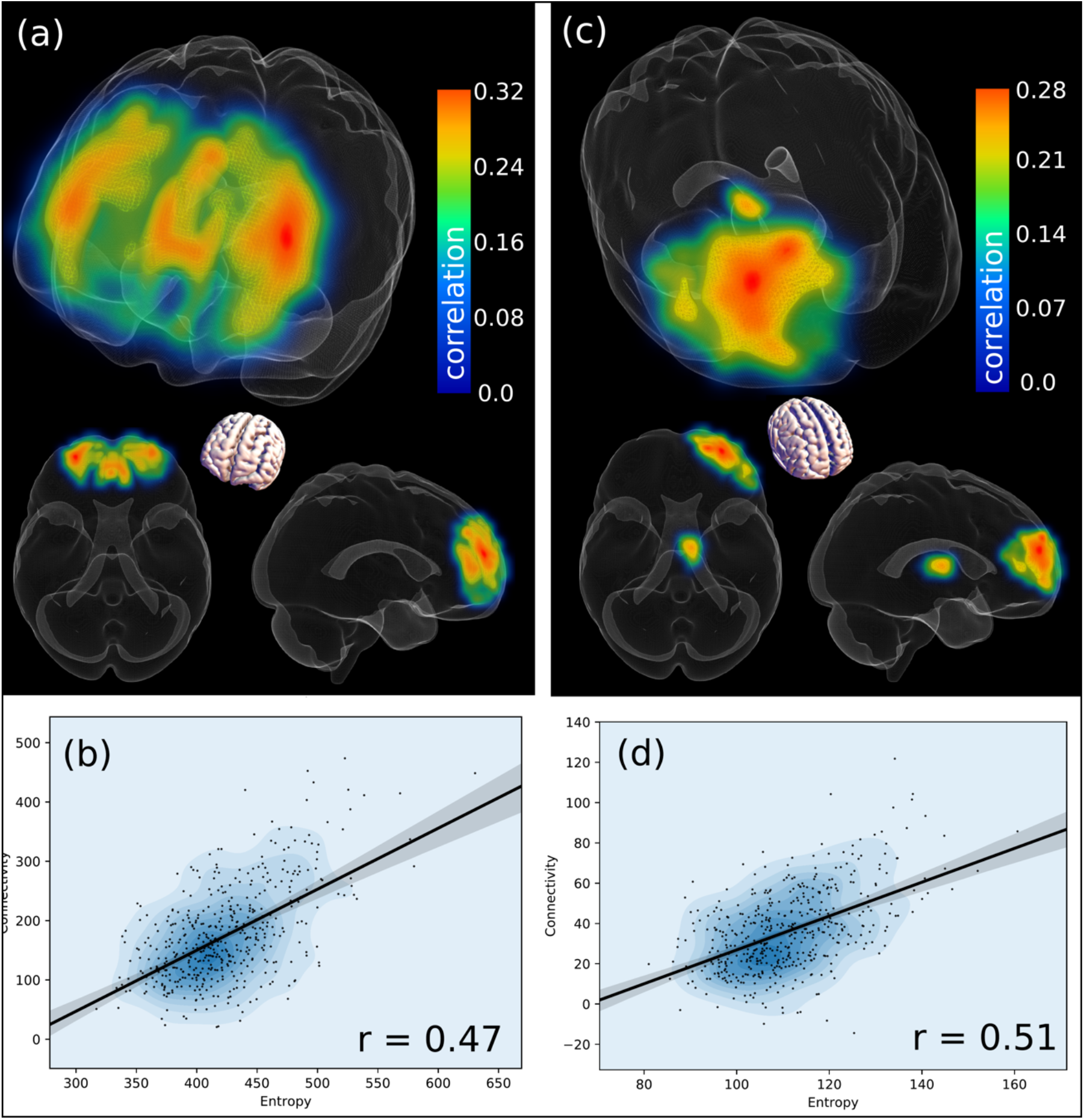
Forceps minor and right ATR voxel-wise entropy correlation with network components summed connectivity. Panels (a) and (c) show the Spearman correlation of entropy voxels with the increased connectivity network component sums (with p<0.001 Bonferroni corrected voxels within wireframe isosurface). (b) and (d) show the correlation and scatterplot with all subjects over summed entropy and summed connectivity on the respective network components.

### 3.4 Relation between functional connectivity and cognition

We proceeded to address the question of how WMH-related functional connectivity changes affected cognitive function. Table 2 details correlations between cognitive tests scores and summed functional connectivity of the significant network components outlined in the previous subsection (Figs. 3, 4, and Supplementary Fig. 1, 2 and 3). The summed functional connectivity on the *reduced* connectivity components correlated negatively with all three AQT tests and to a lesser extent Trailmaking Test A, indicating lower performance (longer test times) with reduced functional connectivity. Moreover, the tract damage metric-associated *increased* connectivity sums correlated positively with AQT and Trailmaking, indicating lower performance also with increased functional connectivity. Both the increased and decreased connectivity are associated with worse cognitive scores, but *reduced* functional connectivity components were mostly related to the AQT Color and AQT Form, whereas *increased* functional connectivity correlated mostly with the executively more demanding AQT Color and Form subpart, as well as Trailmaking Test A.

**Table 2:** Spearman correlations between cognitive tests scores and summed functional connectivity of the WMH related significant network components.

| Tract | AQT Form | AQT Color | AQT<br>Color and Form | Trailmaking<br>Test A |
| --- | --- | --- | --- | --- |
| **Reduced functional connectivity component sum and cognitive test correlations** |  |  |  |  |
| **ATR L** | - | -0.104 (0.025*) | - | - |
| **ATR R** | -0.102 (0.029*) | -0.15 (0.001**) | -0.103 (0.027*) | - |
| **IFOF L** | -0.156 (0.001**) | -0.126 (0.007*) | - | - |
| **IFOF R** | -0.147 (0.002*) | -0.134 (0.004*) | - | - |
| **ILF L** | -0.154 (0.001**) | -0.169 (0.001**) | -0.128 (0.006*) | - |
| **ILF R** | -0.126 (0.007*) | -0.154 (0.001**) | -0.101 (0.03*) | -0.111 (0.03*) |
| **UF L** | -0.102 (0.028*) | - | -0.098 (0.036*) | - |
| **UF R** | - | -0.1 (0.032*) | - | -0.101 (0.047*) |
| **Increased functional connectivity component sum and cognitive test correlations** |  |  |  |  |
| **ATR L** | 0.125 (0.007*) | 0.132 (0.005*) | 0.144 (0.002*) | 0.17 (0.001**) |
| **ATR R** | - | - | 0.101 (0.03*) | 0.125 (0.014*) |
| **IFOF L** | 0.124 (0.008*) | 0.138 (0.003*) | 0.147 (0.002*) | 0.157 (0.002*) |
| **IFOF R** | - | - | 0.103 (0.026*) | 0.138 (0.007*) |
| **UF L** | 0.143 (0.002*) | 0.139 (0.003*) | 0.126 (0.007*) | 0.173 (0.001**) |
| **F minor** | 0.095 (0.041*) | - | - | 0.16 (0.002*) |

### 3.5 Regional WMH and functional connectivity: subgroup with no amyloid pathology

Analyses of the correlations between tract damage functional connectivity in amyloid negative subjects (CSF Aβ42/40>0.1, N = 283) for the tracts of high interest in the above analyses (ATR, IFOF and forceps minor). The results for the amyloid negative subgroup were very similar to those observed using all subjects, indicating a negligible amyloid effect.

## 4. Discussion

Our main findings were that WMHs relate to metrics revealing structural damage and an anatomically divergent pattern of significant functional remodelling. Both increased and decreased functional connectivity were present and both related to worse cognition. Frontally localised connectivity increase was associated with less predictable oxygenation time series, i.e. higher entropy.

### 4.1 Expression of lesion damage in microstructural tissue properties of white matter tracts

A large body of studies shows that, as in the current work, WHMs affect microstructural properties of the tracts as reflected by increased MD and decreased FA (Maniega et al. 2015). For the IFOF (with relatively high lesion volumes) FA did not show a significant intra-extralesional difference and showed a paradoxical increase in ILF lesions (see Fig. 2AB), consistent with previous studies highlighting that MD outperforms FA in differentiating WMHs/normal white matter (Wardlaw et al. 2015,Vernooij et al. 2009). Although intra-relative extralesional FA and MD changes are of the greatest magnitude, we found that extralesional values in subjects with lesions is already significantly altered relative whole tract averages of lesion-free subjects (Fig. 2A, B). This finding, consistent with other research (Tuladhar et al. 2015) implies that while WMHs are hotspots of fibre bundle topological disorder, changes are already detectable in extralesional white matter. This highlights overall tract topology as a potentially early marker of impending lesion formation.

### 4.2 WMH-associated functional connectivity changes

In accordance with previous studies we observed decreased interregional functional connectivity between pairs of grey matter regions with increased lesion load in the connecting tract (Taylor et al. 2016; Langen et al. 2017; Reijmer et al. 2015). The greatest structural-functional associations were found for the IFOF and ATR. Both tracts traverse the lesion probability maxima where the fiber bundles converge in a narrow passage close to the lateral ventricles anterior horns (see Fig. 1, 2 and 3), corresponding to a vascular border zone (Kim et al. 2019). The topology of the damaged segments appears to render them particularly vulnerable with seemingly greater functional disruption. This hypothesis is consistent with a generally more diffuse damage profile (e.g. forceps major) not being associated with significant functional changes.

Additionally, a pattern of *increased* intraregional functional connectivity with growing tract damage emerged, generally confined within the prefrontal grey matter endpoint regions. The increased connectivity appears independent of whether the tracts are association (IFOF, UF), projection (ATR), or commissural tracts (forceps minor), suggesting a phenomenon linked with the frontal cortex. These patterns are consistent with loss of long-range connectivity causing increased short-range connectivity in localized functional subunits and an impaired distributed computing network architecture in the frontal lobe. To our knowledge, this is the first study showing such a possible mechanism in cerebrovascular pathology. Most likely, the pathophysiological process itself affects the long-range connectivity disproportionally more than short-range intracortical connectivity mediated by the short frontal intralobar tracts and U-fibers (Catani et al. 2012). If the diverse inputs to the frontal lobe are disrupted, neuronal activity within the isolated region will exhibit more localized communication patterns, reflected in increased synchronization of oxygenation. Conceptually, this would be very much akin to the white matter disconnection syndrome in the hodological framework by Catani and Ffychte (Catani and Ffytche 2005). Such mechanisms have been demonstrated experimentally, in a hippocampal lesion model in the macaque (Froudist-Walsh et al. 2018).

Attempting to clarify the nature of the increased localized connectivity we employed an information theoretical measure of entropy for discrete time curves (Richman and Moorman 2000). Increased entropy in this context means reduced predictability and increased complexity, whereas reduced entropy means a simpler, more ordered and predictable time-curve. The strong, positive correlations with entropy and the frontal increases in connectivity indicates that these represent increasingly complex, less predictable states. Entropy in neuroscience is not fully understood. Alterations in cerebral function have been associated with both decreases and increases in entropy. Alzheimer’s disease shows regional reduction in entropy (Wang et al. 2017), while in prodromal Alzheimer’s disease (Berron et al. 2020), a localized functional connectivity increase was associated with increased entropy. Normal activation in a sensorimotor fMRI paradigm leads to reduced entropy with BOLD signal increase (Wang et al. 2014), and generally the brains’ task-positive, activated, operating modes involves lower entropy than resting state (Nezafati et al. 2020). Our finding of increased entropy is inconsistent with neurons synchronizing in a regular periodic fashion due to absence of input. Rather, we hypothesize that the increased frontal connectivity could represent suboptimal compensation attempts, which ultimately contribute to cognitive deficits.

Note that the functional connectivity changes cannot only be interpreted in terms of the spatial distribution of WMHs which would predict that the posterior, occipital grey matter regions also should exhibit increased intralobar connectivity with lesion load. The frontal cortex is an richly connected association cortex (Mesulam 2000), chiefly engaged in higher-order processing and may therefore be more susceptible or react differently to reduced inputs via damaged tracts, forcing a remodelling of functional network architecture (Hillary et al. 2014). Also, the location of WMH maxima means that the frontal cortex not only suffers loss of cortico-cortical connections but simultaneously e.g. thalamic, limbic and brainstem reciprocal connections, which may set it aside from other regions and prompt a different response (Jakabek et al. 2018). From a network perspective, the frontal cortex constitutes a hub, i.e. a highly connected region with network properties that fulfil a crucial role in the overall network organization, also highlighting this region as particularly susceptible to damage (van den Heuvel and Sporns 2011), verified through simulations and with experimental support (Froudist-Walsh et al. 2018).

### 4.3 Cognitive consequences of structural-functional alterations

Both increase and decrease in functional connectivity were associated with worse cognition. Longer test duration for all three AQT tests (indicating worse performance) correlated with the reduced connectivity components, in concordance with findings of previous studies (Duering et al. 2014). However, the WMH-associated connectivity increase correlated with reduced performance in AQT (particularly Color and Form) and trailmaking Test A. If we consider increased connectivity as a compensatory mechanism to counterbalance long-range communication failure, it appears to represent a suboptimal or failed attempts, and should therefore likely be a consequence of relatively severe disruptions.

### 4.4 Amyloid pathology, WMH and functional connectivity

Results for the subgroup with no amyloid pathology were very similar to the ones including all subjects, suggesting an amyloid-independent effect of WMH on functional connectivity and cognition. This is in line with several other studies that have shown that these processes contribute independently to the compromised functional connectivity (Marchant et al 2011, Taylor et al 2016).

### 4.5 Methodological considerations

There are several methodological aspects to consider in this study. Firstly, the tract damage associated functional changes and their relation to cognition are not very strong. Given the complex relationship between structural connectivity, functional connectivity and cognition, this is perhaps to be expected but also indicative of relatively weak effects. Second, the summed and weighted tract damage metric does not really compare with detailed evaluation of fiber tracking on an individual basis, but as fiber tracking is fraught with methodological difficulties (Maier-Hein et al. 2017). Therefore, we favoured a simpler approach. However, with the large sample size of our study we are still able to provide clear patterns associated with structural communication disruption. Additionally, we chose to apply a combined measure of volume and DTI metrics within the lesion to better capture lesion severity. A large-sized lesion can sometimes be less damaging to the underlying tissue, whereas a small lesion can be a so-called black-hole with actual fibre tract disruption, which is expected to have a larger histopathological severity (Gouw et al. 2011), better accounted for by the probability-weighted intralesional sum of MD. Finally, we only considered major white matter tracts with clearly definable endpoints, and only regions directly (and not indirectly) connected via the white matter tracts.

## 5.0 Conclusion

In the present work we demonstrate that the effect on structural connectivity of WHMs are associated with different functional connectivity responses depending on anatomy. There is an association with frontally localized intralobar increase and simultaneously interlobar decrease of connectivity. We speculate that anatomical divergent response is linked to the connectivity anatomy and functionality of the prefrontal lobe. The frontal increase in connectivity is related to worse cognition, and to higher entropy, the latter currently understood as an index of reduced efficiency. Therefore we propose that this increase of functional connectivity represents failed compensatory attempt.

## Data Availability

Anonymized data will be shared by request from a qualified academic investigator for the sole purpose of replicating procedures and results presented in the article if data transfer is in agreement with EU legislation on the general data protection regulation and decisions by the Ethical Review Board of Sweden and Region Skane, which should be regulated in a material transfer agreement.

## Acknowledgements

We express our gratitude toward all participating subjects, their next of kin, to all personnel of the Swedish BioFINDER study, and the funding bodies.

## Abbreviations

AQT: A Quick Test for Cognitive Speed
ATR: anterior thalamic radiation
Cing-hipp: cingulum hippocampal part
Cing-cing: cingulum cingulate gyrus part
CN: cognitively normal
CST: cortico-spinal tract
FA: fractional anisotropy
IFOF: inferior fronto-occipital fasciculus
ILF: inferior longitudinal fasciculus
L/R: (left and right hemisphere)
MCI: mild cognitive impairment
MD: mean diffusivity
SLF: superior longitudinal fasciculus
UF: uncinate fasciculus
WMHs: white matter hyperintensities

